# Lockdown may partially halt the spread of 2019 novel coronavirus in Hubei province, China

**DOI:** 10.1101/2020.02.11.20022236

**Authors:** Mingwang Shen, Zhihang Peng, Yuming Guo, Yanni Xiao, Lei Zhang

## Abstract

We present a timely evaluation of the impact of lockdown on the 2019-nCov epidemic in Hubei province, China. The implementation appears to be effective in reducing about 60% of new infections and deaths, and its effect also appears to be sustainable even after its removal. Delaying its implementation reduces its effectiveness. However, the direct economic cost of such a lockdown remains to be seen and whether the model is replicable in other Chinese regions remains a matter of further investigation.

---

On 31^st^ December 2019, a pneumonia case of unknown etiology from Wuhan City, Hubei Province, China was reported to the World Health Organization (WHO), which was later confirmed as a novel coronavirus and named ‘2019-nCoV’ [1] (renamed as ‘COVID-19’ after one month [2]). As of 11th February 2020, 44,653 confirmed cases and 1,113 deaths have been reported in China, and approximately 75% of (33,366) confirmed cases and >95% (1,068) of deaths were from Hubei province [3]. Preliminary studies have suggested that 2019-nCoV is much more infectious but less deadly [4,5] than the Severe Acute Respiratory Syndrome (SARS) and the Middle East Respiratory Syndrome (MERS). 2019-nCoV is capable of human-to-human transmission even during its asymptomatic incubation period (3-7 days [6]). On 23^rd^ January, the Chinese government initiated an unprecedented public health measure to confine the epidemic by locking down 13 cities in Hubei province, preventing movement of its population by terminating most forms of transportation [7,8]. We provide a brief analysis of the population impact of the lockdown on the 2019-nCoV epidemic by investigating various scenarios of ‘delayed’ or ‘no lockdown’ based on a mathematical model (**Appendix**).

Overall, the lockdown may partially halt the spread of the epidemic in Hubei province. In the status quo, as the lockdown remains effective when this article is written, we assume the lockdown will last for three weeks based on the official schedule of the working day. The lockdown has effectively shut down most forms of transportation, both public and private, we estimate the number of person-to-person contacts per day among its residents has reduced from thirteen to ten. The percentage of facial mask usage in public space has also drastically increased to almost 100% in Hubei province during the lockdown. However, the increase may be due to people’s perception of the severity of the epidemic, rather than the lockdown itself. After the lockdown period of 21 days, we assumed the person-to-person contact rate will return to normal but mask usage will retain. With these assumptions, we forecast that the number of confirmed 2019-nCoV cases will reach its peak of 12,143 (5,872-19,852) individuals at 27 (23-36) days (i.e., 17 Feb [14-27 Feb]) after the lockdown initiation (Figure 1c). The trend will then gradually decline and the cumulative number of infections and death cases in Hubei province will reach 128,960 (39,362-218,560) and 4,381 (1,667-7,095) at the end of the epidemic (Figure 1a), corresponding to a fatality rate of 3.44% (3.19-3.70%). On contrast, if the lockdown has been delayed for seven days, we may observe a larger number of confirmed cases at peak (18,095 [8,618-28,274]), the total infections (169,030 [53,238-284,820]) and deaths (5,905 [2,254-9,556]). Further, if the delay has been 14 days, the epidemic will peak at 25,194 (11,634-39,667) confirmed cases and the total infections and deaths will reach 210,400 (66,785-354,020) and 7,451 (2,784-12,118), respectively. In the case where no lockdown was in place, the epidemic will peak at 36,544 (13,919-61,243) confirmed cases and the total infections and death cases will reach 348,750 (84,140-613,360) and 11,076 (3,408-18,744), respectively. This suggests that the current lockdown strategy may eventually prevent 219,790 (44,773-394,800) 2019-nCoV infections and 6,695 (1,741-11,650) deaths, that is, 62.53% (59.98-65.08%) of infections and 59.91% (56.87-62.95%) deaths, compared with no lockdown. We observed a small re-bounce in the number of latent individuals when the 21-day lockdown is lifted (Figure 1b). However, this re-bounce does not fully account for the return influx of the ‘travel rush’ and cannot determine if a second epidemic will follow.

**Figure 1.**
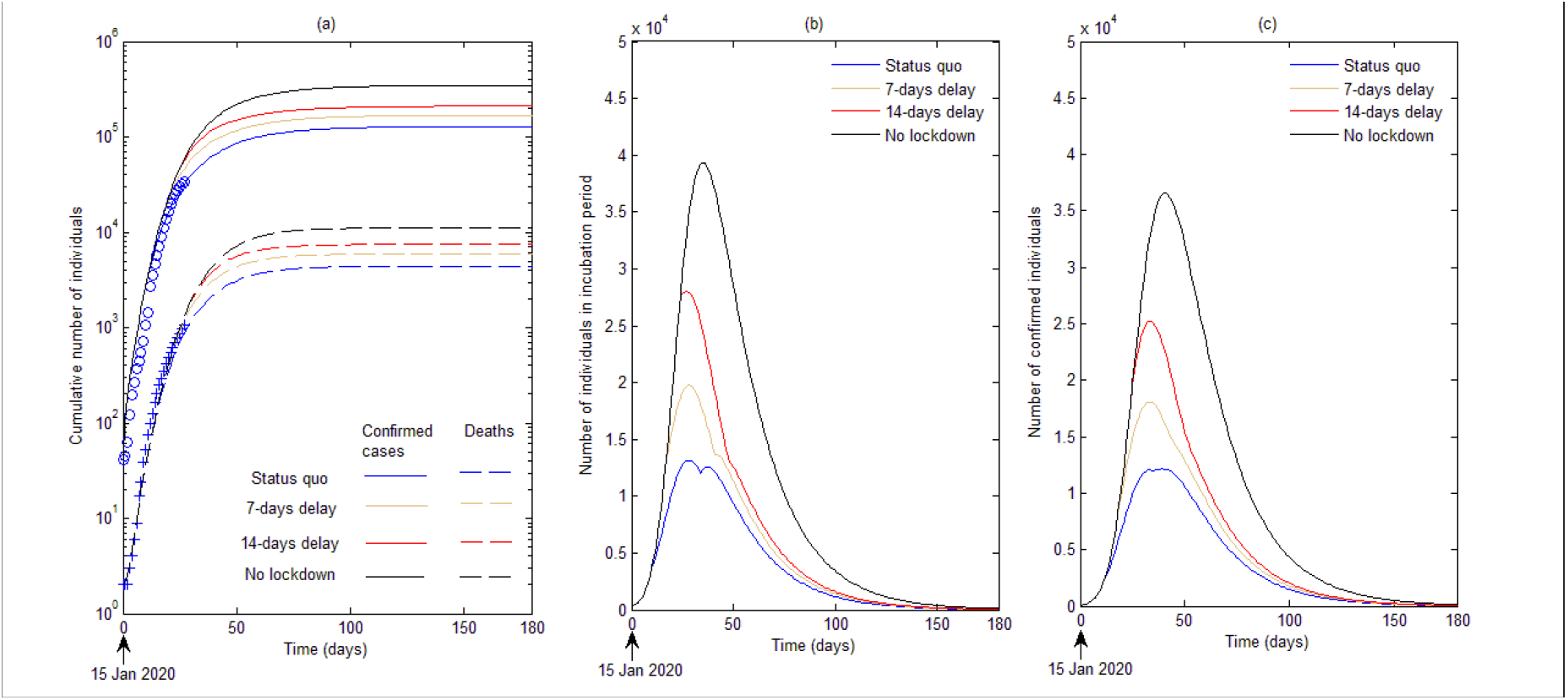
(a) Calibration of reported confirmed 2019-nCoV cases and deaths in Hubei province, China; (b) number of individuals in incubation period and (c) confirmed individuals over time for four scenarios: status quo (lockdown during 23^rd^ January to 12^th^ February 2020), 7-days delay, 14-days delay, and ‘no lockdown’.

We present a timely evaluation of the impact of ‘lockdown’ on the 2019-nCov epidemic in Hubei province, China. The implementation appears to be effective in reducing about 60% of new infections and deaths, and its effect also appears to be sustainable even after its removal. Delaying its implementation reduces its effectiveness. However, the direct economic cost of such a lockdown remains to be seen and whether the model is replicable in other Chinese regions remains a matter of further investigation.

All authors declare that they have no competing interests.

## Data Availability

Data was attached in the Appendix.

## Acknowledgments

This work was supported by the National Natural Science Foundation of China (grant numbers: 8191101420(LZ), 11801435 (MS), 11631012 (YX), 81673275(ZP), 91846302(ZP)); Thousand Talents Plan Professorship for Young Scholars (grant number 3111500001); Xi’An Jiaotong University Young Talent Support Program; China Postdoctoral Science Foundation (grant number 2018M631134); the Fundamental Research Funds for the Central Universities (grant number xjh012019055); Natural Science Basic Research Program of Shaanxi Province (Grant number: 2019JQ-187); the National S&T Major Project Foundation of China (2018ZX10715002-004, 2018ZX10713001-001) and the Priority Academic Program Development of Jiangsu Higher Education Institutions (PAPD). YG was supported by Career Development Fellowships of the Australian National Health and Medical Research Council (numbers APP1107107 and APP1163693).

## Authors’ contributions

M.S., Z.P., Y.G., Y.X., and L.Z. conceived and designed the study. M.S. analyzed the data, carried out the analysis and performed numerical simulations. M.S. wrote the first draft of the manuscript. M.S., Z.P., Y.G., Y.X., and L.Z. contributed to writing the paper and agreed with manuscript results and conclusions.

